# Advanced HIV disease and treatment-related adverse drug reactions among people living with HIV receiving antiretroviral therapy in Tanzania: a multicenter cross-sectional study

**DOI:** 10.64898/2026.05.30.26354502

**Authors:** Ritah F. Mutagonda, Wema A Kibanga, Wigilya P. Mikomangwa, Appolinary AR. Kamuhabwa

## Abstract

**Background:** Advanced HIV disease (AHD) remains a major contributor to HIV-related morbidity and mortality despite widespread antiretroviral therapy (ART) access in sub-Saharan Africa. Although treatment-related adverse drug reactions (ADRs) may compromise treatment outcomes, evidence on the relationship between AHD and ADR occurrence remains limited. This study aimed to determine the prevalence and identify factors associated with AHD, characterize treatment-related ADR and assess the association between AHD and ADR occurrence among people living with HIV receiving ART in Dar es Salaam, Tanzania.

**Methods:** We conducted a multicenter cross-sectional study among 1,513 people living with HIV receiving ART at selected HIV care and treatment clinics in Dar es Salaam, TanzaniaFor this adolescent/adult cohort, AHD was operationally defined as WHO clinical stage III/IV disease and/or baseline CD4 count <200 cells/mm^3^. Treatment-related ADRs were defined as participant-reported and/or clinically documented ART-related adverse events identified during routine HIV care, including both current and retrospectively reported events. Modified Poisson regression with robust standard errors was used to estimate crude and adjusted risk ratios (RRs) with 95% confidence intervals (CIs).

**Results:** Among 1,508 participants with sufficient information for classification, 961 (63.7%) had AHD. Factors independently associated with AHD included age ≥50 years (aRR 1.10, 95% CI 1.01–1.20), underweight nutritional status (aRR 1.17, 95% CI 1.00–1.35), and concomitant medication use (aRR 1.19, 95% CI 1.03–1.37), while DTG-based ART was associated with lower AHD prevalence (aRR 0.78, 95% CI 0.68–0.90). Overall, 569 participants (38.0%) reported at least one ADR. Composite AHD was not independently associated with ADR occurrence (aRR 0.95, 95% CI 0.82–1.11), but baseline CD4 <200 cells/mm^3^ was associated with increased ADR risk (aRR 1.20, 95% CI 1.02–1.41). Comorbidity (aRR 1.66, 95% CI 1.42–1.93) was the strongest correlate of ADR occurrence.

**Conclusion:** AHD remains highly prevalent among people living with HIV receiving ART in Tanzania. While composite AHD was not independently associated with ADR occurrence, severe immunosuppression, comorbidity burden, and concomitant medication exposure were associated with increased ADR risk. These findings suggest that immunologic severity and broader clinical complexity may be more informative predictors of ART-related toxicity than composite syndromic AHD classification alone. Strengthened early diagnosis, differentiated advanced HIV care, integrated pharmacovigilance strategies, and routine medication risk assessment are needed.

## Introduction

Human immunodeficiency virus (HIV) infection remains a major global public health challenge despite substantial advances in prevention, diagnosis, and treatment. In 2023, an estimated 39.9 million people were living with HIV globally, with sub-Saharan Africa continuing to bear the greatest burden of disease. Tanzania remains among the high HIV burden countries in the region, with approximately 1.5 million people living with HIV and an adult HIV prevalence of approximately 4.4% [1]. The scale-up of antiretroviral therapy (ART), particularly following adoption of universal treatment policies, has substantially reduced HIV-related morbidity and mortality and transformed HIV infection into a manageable chronic condition for many individuals [2,3]. However, despite these gains, advanced HIV disease (AHD) remains an important unresolved clinical and public health challenge.

The World Health Organization (WHO) defines AHD as WHO clinical stage III or IV disease, CD4 cell count below 200 cells/mm^3^, or HIV infection in children younger than five years [4]. For adolescents and adults, AHD is primarily identified using severe immunosuppression and/or advanced clinical staging criteria. Although universal ART access was expected to reduce late HIV presentation and severe immunosuppression, recent evidence suggests that AHD remains common in routine HIV care settings. A recent global review reported that approximately one-third of individuals presenting to outpatient HIV care settings had AHD, while prevalence was substantially higher in hospitalized populations [5]. Similarly, a recent systematic review and meta-analysis from sub-Saharan Africa reported a pooled AHD prevalence of 58.7%, underscoring the continuing burden of advanced disease in the region [6]. Tanzanian data reflect similar concerns, with cohort evidence showing that more than half of individuals entering HIV care continue to present with advanced disease despite implementation of Treat All strategies [7].

The persistence of AHD in the ART era reflects multiple intersecting challenges, including delayed HIV diagnosis, late linkage to care, treatment interruption, poor retention, virologic failure, and inadequate recognition of immunologic deterioration among ART-experienced patients [8,9,10]. Importantly, the burden of AHD in contemporary HIV programs increasingly reflects not only late presentation to care, but also ongoing clinical complexity among treatment-experienced individuals. While ART programs have increasingly shifted focus toward viral load monitoring, CD4 testing remains clinically important for identifying individuals requiring differentiated AHD care, including opportunistic infection screening, prophylaxis, and intensified clinical monitoring [4].

Although ART has substantially improved survival and long-term HIV outcomes, treatment-related adverse drug reactions (ADRs) remain an important barrier to optimal HIV care [11–13]. ART-related ADRs range from relatively mild tolerability-related events such as nausea, dizziness, and dermatologic reactions to more serious neurologic, renal, hepatic, hematologic, and hypersensitivity toxicities [11]. These adverse events may impair treatment adherence, necessitate regimen modification, increase healthcare utilization, and compromise treatment success [12,13]. In routine HIV care settings, ADR burden may be further complicated by multimorbidity, polypharmacy, and cumulative exposure to multiple ART regimens over time.

A biologically plausible relationship exists between AHD and treatment-related ADRs. Individuals with severe immunosuppression may be more vulnerable to toxicity because of impaired physiologic reserve, chronic systemic inflammation, malnutrition, organ dysfunction, opportunistic co-infections, and exposure to multiple concomitant medications, including prophylactic and therapeutic agents [11,13,14]. Polypharmacy in clinically complex patients may further increase the risk of cumulative toxicity and drug–drug interactions [13]. However, the mechanisms linking advanced disease severity and ART-related toxicity are likely heterogeneous. Severe immunosuppression, advanced clinical syndromes, comorbidity burden, and concomitant medication exposure may contribute differently to ADR susceptibility.

Understanding the relationship between AHD and ART-related toxicity has important implications for clinical risk stratification, differentiated HIV service delivery, and pharmacovigilance strengthening in resource-limited settings where both advanced disease and treatment complexity remain common. This study therefore aimed to determine the prevalence of AHD, identify factors associated with AHD, characterize treatment-related ADRs, and assess the association between AHD and ADR occurrence among people living with HIV receiving antiretroviral therapy in Dar es Salaam, Tanzania.

## Methods

### Study design and setting

A multicenter cross-sectional analytical study was conducted among people living with HIV receiving ART in Tanzania from May 2022 to June 2023. The study was conducted in selected HIV care and treatment clinics (CTCs) in Dar es Salaam, Tanzania, including Muhimbili National Hospital (MNH), Mwananyamala Regional Referral Hospital, Temeke Regional Referral Hospital, Amana Regional Referral Hospital, and Mnazi Mmoja Hospital. These facilities are major public HIV treatment centers providing routine HIV care, ART initiation, treatment monitoring, and pharmacovigilance services under the Tanzanian national HIV treatment program. During the study period, Tanzania had adopted universal ART initiation strategies and was transitioning toward widespread implementation of DTG-based regimens as preferred first-line treatment.

### Study population and participant recruitment

The study population comprised adolescents and adults with confirmed HIV infection receiving ART at participating HIV care and treatment clinics. Participants were recruited consecutively during routine clinic attendance at the selected study sites using standardized study procedures. Data were collected through direct participant interviews and abstraction of relevant clinical and treatment information from patient medical records.

Eligible participants were individuals with confirmed HIV infection receiving ART during the study period who had sufficient socio-demographic, clinical, and treatment information available for analysis. Participants with incomplete information required for specific analyses were excluded from those analyses. A total of 1,513 participants were enrolled.

### Data collection and study variables

Data were collected using standardized structured questionnaires and clinical record abstraction forms. Information obtained included socio-demographic characteristics, clinical parameters, HIV treatment history, comorbidities, concomitant medication use, anthropometric measures, and ART-related ADRs. Socio-demographic variables included age, sex, marital status, educational attainment, and employment status. Age was categorized as <18 years, 18–34 years, 35–49 years, and ≥50 years, while continuous age was summarized using medians and interquartile ranges (IQRs). Clinical variables included body mass index (BMI), WHO HIV clinical stage, baseline CD4 cell count, presence of comorbidities, and AHD status. BMI was categorized as underweight (<18.5 kg/m^2^), normal (18.5–24.9 kg/m^2^), overweight (25.0–29.9 kg/m^2^), and obese (≥30.0 kg/m^2^). Baseline CD4 cell count was categorized as <200 cells/mm^3^, 200–499 cells/mm^3^, and ≥500 cells/mm^3^.

Treatment-related variables included ART regimen category and concomitant medication use. ART regimens were categorized as DTG-based or non-DTG-based regimens. Concomitant medication use was defined as use of non-ART medications, including prophylactic and therapeutic agents, during HIV treatment. The primary exposure variable was AHD. For this adolescent/adult cohort, AHD was operationally defined as WHO clinical stage III/IV disease and/or baseline CD4 count <200 cells/mm^3^. The primary outcome variable was ART-related ADR occurrence, defined as participant-reported and/or clinically documented adverse events attributed to antiretroviral therapy during routine HIV care. ADR ascertainment included both symptoms present at study enrollment and retrospectively reported prior ART-related adverse events documented in clinical records or recalled during participant interviews. WHO clinical staging reflected routinely documented clinical status at or around the time of ADR assessment. No formal ADR causality assessment scale or severity grading system was applied. For sensitivity analyses, the individual components of AHD were examined separately, including WHO clinical stage III/IV disease and baseline CD4 count <200 cells/mm^3^.

### Statistical analysis

Data were cleaned, coded, and analyzed using standard statistical procedures. Categorical variables were summarized using frequencies and percentages. Continuous variables were assessed for distributional normality and, because principal continuous variables were non-normally distributed, were summarized using medians and interquartile ranges (IQRs). The prevalence of AHD was calculated as the proportion of participants meeting the operational WHO-based AHD definition among participants with sufficient information for classification. Similarly, ADR prevalence was calculated as the proportion of participants reporting at least one ART-related ADR. Because AHD and ADR occurrence were relatively common outcomes, modified Poisson regression with robust standard errors was used instead of logistic regression to estimate crude and adjusted risk ratios (RRs) with corresponding 95% confidence intervals (CIs).

Two main regression models were constructed. The first model evaluated factors associated with AHD, with AHD status as the binary outcome variable. Covariates considered included age group, sex, BMI category, ART regimen category, comorbidity status, and concomitant medication use. These covariates were selected a priori based on biological plausibility, clinical relevance, and existing HIV pharmacovigilance literature. The second model assessed factors associated with ART-related ADR occurrence, with ADR occurrence as the binary outcome variable and AHD as the principal exposure variable. This model was adjusted for age group, sex, BMI category, ART regimen category, comorbidity status, and concomitant medication use.

Sensitivity analyses were conducted by replacing the composite AHD variable with its individual components (WHO stage III/IV disease and baseline CD4 count <200 cells/mm^3^) to evaluate whether specific markers of disease severity showed differential associations with ADR occurrence. To minimize collinearity between composite AHD and its individual clinical and immunologic components, these variables were examined separately in sensitivity analyses. A p-value of <0.05 was considered statistically significant. Missing observations were reported descriptively in baseline characteristic tables. Regression analyses were conducted using complete-case observations for variables included in each model, resulting in variable-specific sample sizes depending on data completeness.

### Ethical considerations

Ethical approval for the study was obtained from the Institutional Review Board of Muhimbili University of Health and Allied Sciences (MUHAS) (Approval No. MUHAS-REC-03-2022-1050) prior to study implementation. Written informed consent was obtained from all adult participants before enrollment. For participants aged below 18 years, written assent was obtained from the participant together with written consent from parents or legal guardians in accordance with ethical requirements. Participant confidentiality was maintained through anonymization of study data and restricted access to identifiable information.

## Results

### Participant characteristics

A total of 1,513 people living with HIV receiving antiretroviral therapy (ART) were included in the analysis. The median age was 45 years (interquartile range [IQR]: 36–52). Most participants were aged 35–49 years (36.7%), followed by those aged ≥50 years (43.2%), while 67 participants (4.5%) were aged below 18 years. Females constituted the majority of participants (68.7%). The median body mass index (BMI) was 25.0 kg/m^2^ (IQR: 21.4–29.0). Among participants with available anthropometric data, 9.3% were underweight, 35.6% had normal BMI, 25.4% were overweight, and 16.4% were obese.

Clinically, 823 participants (54.4%) had documented WHO clinical stage III or IV disease during routine HIV care assessment. The median baseline CD4 cell count was 361 cells/mm^3^ (IQR: 173– 572), and 364 participants (24.1%) had baseline CD4 counts below 200 cells/mm^3^. Most participants were receiving DTG-based ART regimens (92.1%), while 7.2% were receiving non-DTG regimens. Comorbid conditions were documented in 26.6% of participants, and 80.8% were using at least one concomitant non-ART medication, including prophylactic or therapeutic agents. Overall, 569 participants (38.0%) reported at least one ART-related ADR (Table 1).

**Table 1.**
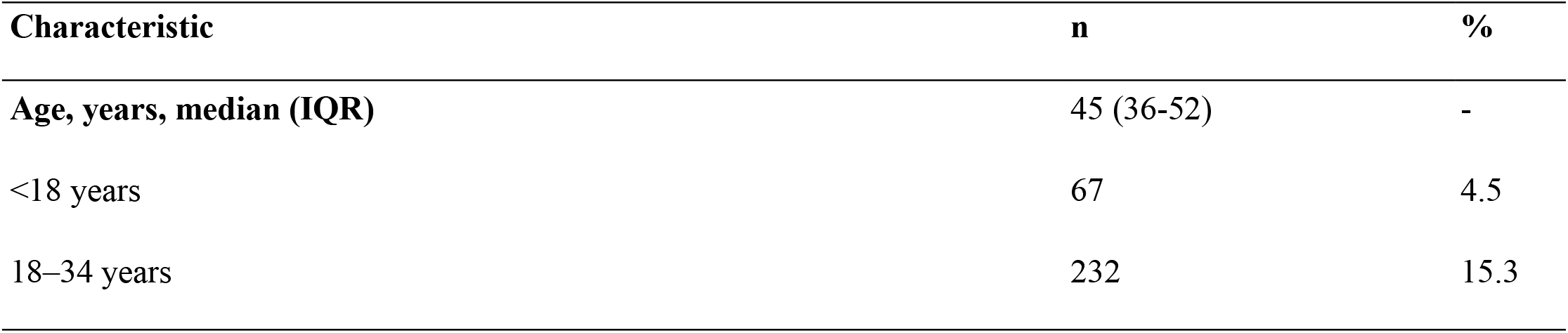
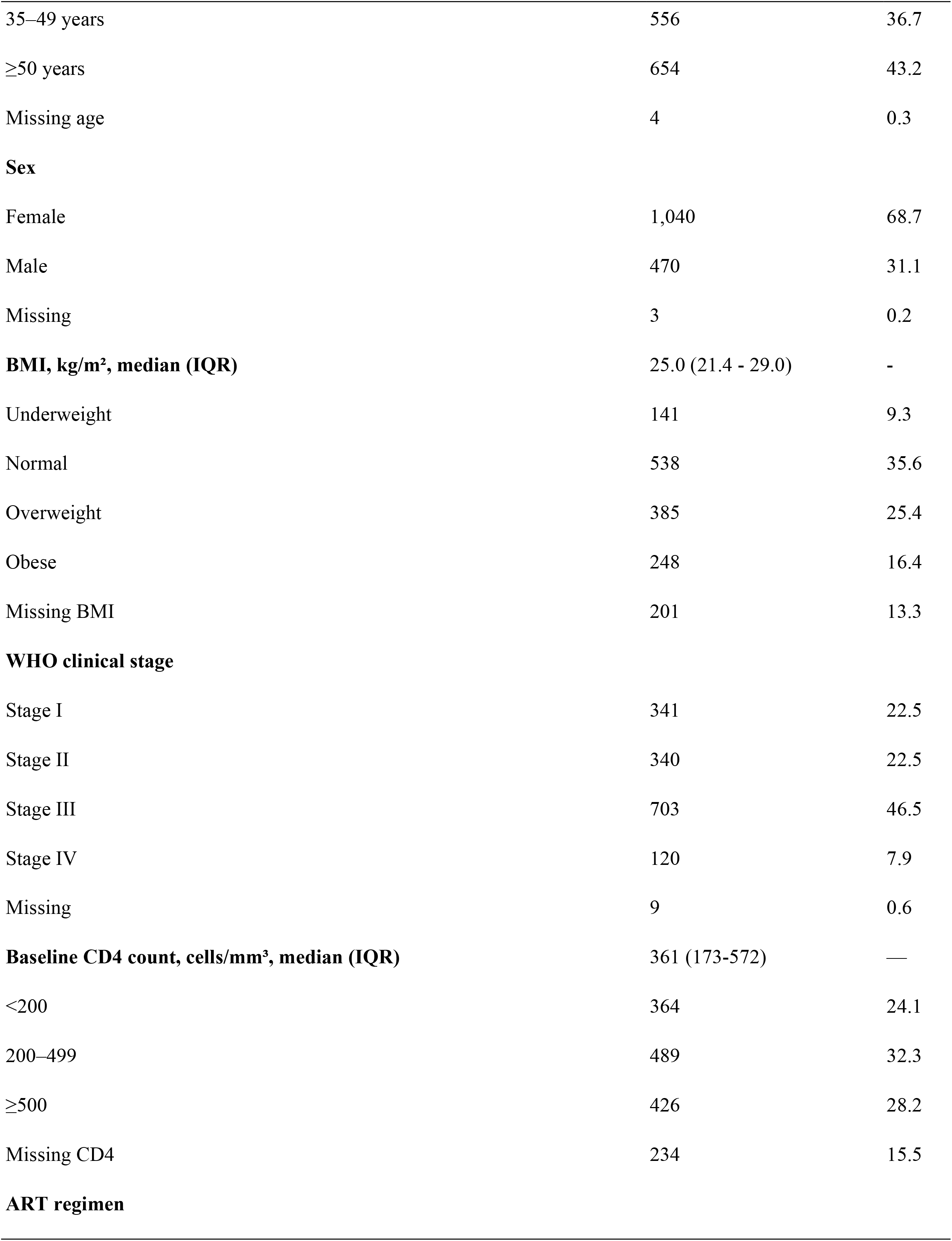
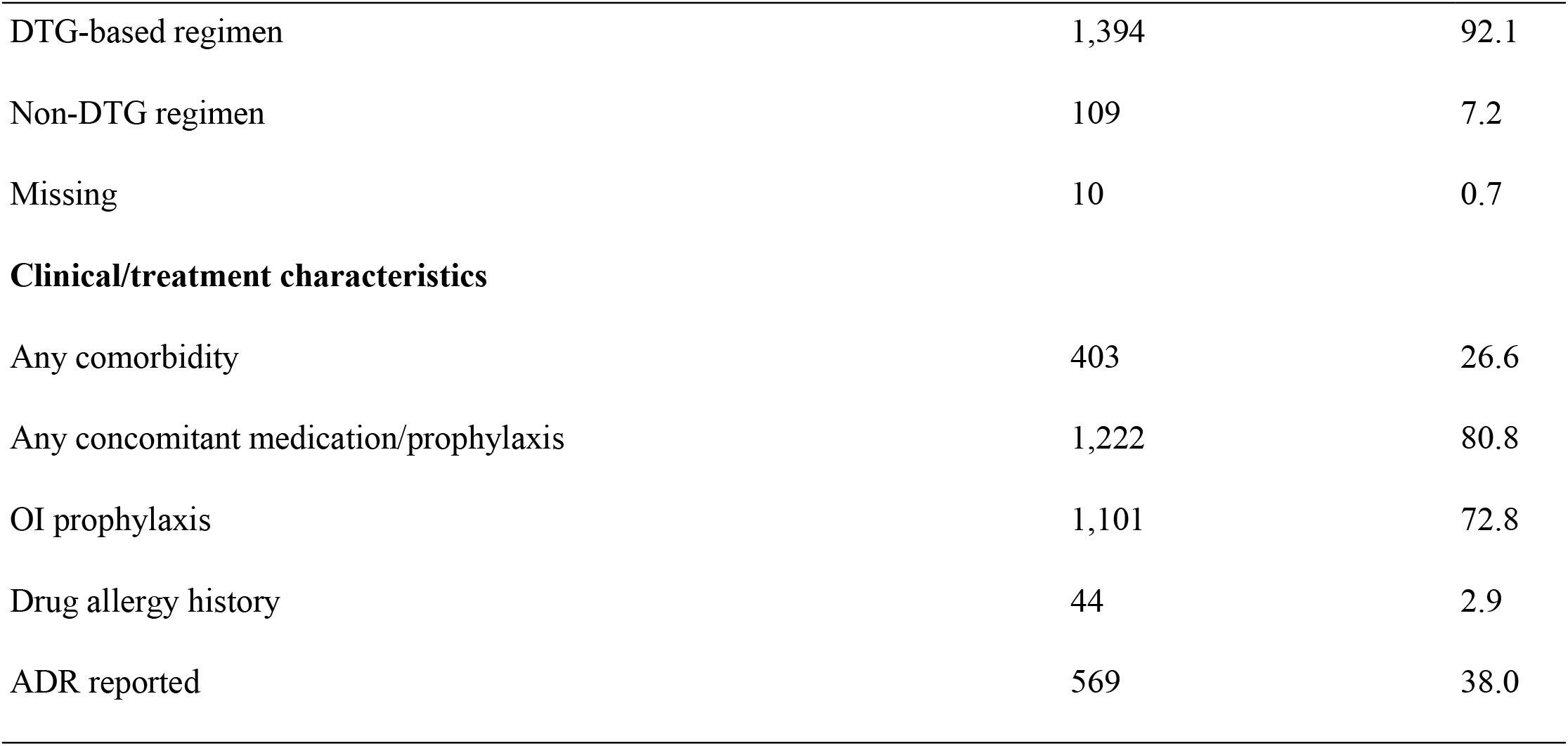
Baseline characteristics of study participants (n=1,513)

| Characteristic | n | % |
| --- | --- | --- |
| Age, years, median (IQR) | 45 (36-52) | - |
| <18 years | 67 | 4.5 |
| 18–34 years | 232 | 15.3 |

| **Characteristic** | **n** | **%** |
| --- | --- | --- |
| 35–49 years | 556 | 36.7 |
| ≥50 years | 654 | 43.2 |
| Missing age | 4 | 0.3 |
| **Sex** |  |  |
| Female | 1,040 | 68.7 |
| Male | 470 | 31.1 |
| Missing | 3 | 0.2 |
| **BMI, kg/m<sup>2</sup>, median (IQR)** | 25.0 (21.4 - 29.0) | - |
| Underweight | 141 | 9.3 |
| Normal | 538 | 35.6 |
| Overweight | 385 | 25.4 |
| Obese | 248 | 16.4 |
| Missing BMI | 201 | 13.3 |
| **WHO clinical stage** |  |  |
| Stage I | 341 | 22.5 |
| Stage II | 340 | 22.5 |
| Stage III | 703 | 46.5 |
| Stage IV | 120 | 7.9 |
| Missing | 9 | 0.6 |
| **Baseline CD4 count, cells/mm<sup>3</sup>, median (IQR)** | 361 (173-572) | — |
| <200 | 364 | 24.1 |
| 200–499 | 489 | 32.3 |
| ≥500 | 426 | 28.2 |
| Missing CD4 | 234 | 15.5 |
| **ART regimen** |  |  |
| DTG-based regimen | 1,394 | 92.1 |
| Non-DTG regimen | 109 | 7.2 |
| Missing | 10 | 0.7 |
| **Clinical/treatment characteristics** |  |  |
| Any comorbidity | 403 | 26.6 |
| Any concomitant medication/prophylaxis | 1,222 | 80.8 |
| OI prophylaxis | 1,101 | 72.8 |
| Drug allergy history | 44 | 2.9 |
| ADR reported | 569 | 38.0 |

### Burden of advanced HIV disease

Using the operational WHO-based definition applied in this adolescent/adult cohort (WHO clinical stage III/IV disease and/or baseline CD4 count <200 cells/mm^3^), 961 of 1,508 participants with sufficient information for classification had AHD, corresponding to a prevalence of 63.7%. Among participants classified as having AHD, 823 (54.6%) met WHO clinical staging criteria and 364 (24.1%) had severe immunosuppression defined by baseline CD4 count below 200 cells/mm^3^.

### Factors associated with advanced HIV disease

In univariate analysis, younger age (18–34 years), male sex, underweight nutritional status, presence of comorbidity, concomitant medication use, and ART regimen category were associated with AHD. In multivariable modified Poisson regression analysis, participants aged 18–34 years had significantly lower prevalence of AHD compared with those aged 35–49 years (adjusted RR [aRR] 0.74, 95% CI 0.63–0.87; p<0.001), whereas participants aged ≥50 years had significantly higher prevalence (aRR 1.10, 95% CI 1.01–1.20; p=0.034). Underweight participants showed modestly higher prevalence of AHD compared with those with normal BMI (aRR 1.17, 95% CI 1.00–1.35; p=0.046). Participants receiving DTG-based ART regimens had lower prevalence of AHD compared with those receiving non-DTG regimens (aRR 0.78, 95% CI 0.68–0.90; p<0.001).

Concomitant medication use was independently associated with higher prevalence of AHD (aRR 1.19, 95% CI 1.03–1.37; p=0.015) (Table 2).

**Table 2.** Univariate and multivariable modified Poisson regression analysis of factors associated with advanced HIV disease.

| Variable | Crude RR (95% CI) | p-value | Adjusted RR (95% CI) | p-value |
| --- | --- | --- | --- | --- |
| **Age group (years)** |  |  |  |  |
| 35–49 (Reference) | 1.00 | — | 1.00 | — |
| <18 | 0.89 (0.71–1.12) | 0.322 | 0.83 (0.65–1.07) | 0.157 |
| 18–34 | 0.79 (0.68–0.92) | 0.003 | 0.74 (0.63–0.87) | <0.001 |
| ≥50 | 1.12 (1.03–1.22) | 0.011 | 1.10 (1.01–1.20) | 0.034 |
| **Sex** |  |  |  |  |
| Female (Reference) | 1.00 | — | 1.00 | — |
| Male | 1.13 (1.04–1.23) | 0.005 | 1.09 (1.00–1.19) | 0.051 |
| **BMI category** |  |  |  |  |
| Normal BMI (Reference) | 1.00 | — | 1.00 | — |
| Underweight | 1.28 (1.11–1.47) | 0.001 | 1.17 (1.00–1.35) | 0.046 |
| Overweight | 1.04 (0.94–1.15) | 0.423 | 1.06 (0.96–1.18) | 0.245 |
| Obese | 1.09 (0.98–1.22) | 0.122 | 1.10 (0.98–1.23) | 0.104 |
| **ART regimen** |  |  |  |  |
| Non-DTG (Reference) | 1.00 | — | 1.00 | — |
| DTG-based | 0.72 (0.64–0.81) | <0.001 | 0.78 (0.68–0.90) | <0.001 |
| **Presence of comorbidity** |  |  |  |  |
| None (Reference) | 1.00 | — | 1.00 | — |
| Yes | 1.12 (1.03–1.23) | 0.012 | 1.05 (0.96–1.15) | 0.300 |
| **Concomitant medication/OI prophylaxis** |  |  |  |  |
| None (Reference) | 1.00 | — | 1.00 | — |
| Yes | 1.26 (1.11–1.43) | <0.001 | 1.19 (1.03–1.37) | 0.015 |

### Burden and pattern of treatment-related ADRs

Overall, 569 of 1,497 participants with available ADR outcome data (38.0%) reported at least one ART-related ADR. The most frequently reported ADRs were itching/pruritus (17.6%), dizziness (14.5%), nausea (6.8%), skin rash (5.1%), and drowsiness or sleep disturbance (4.3%) (Figure 1).

**Figure 1.**
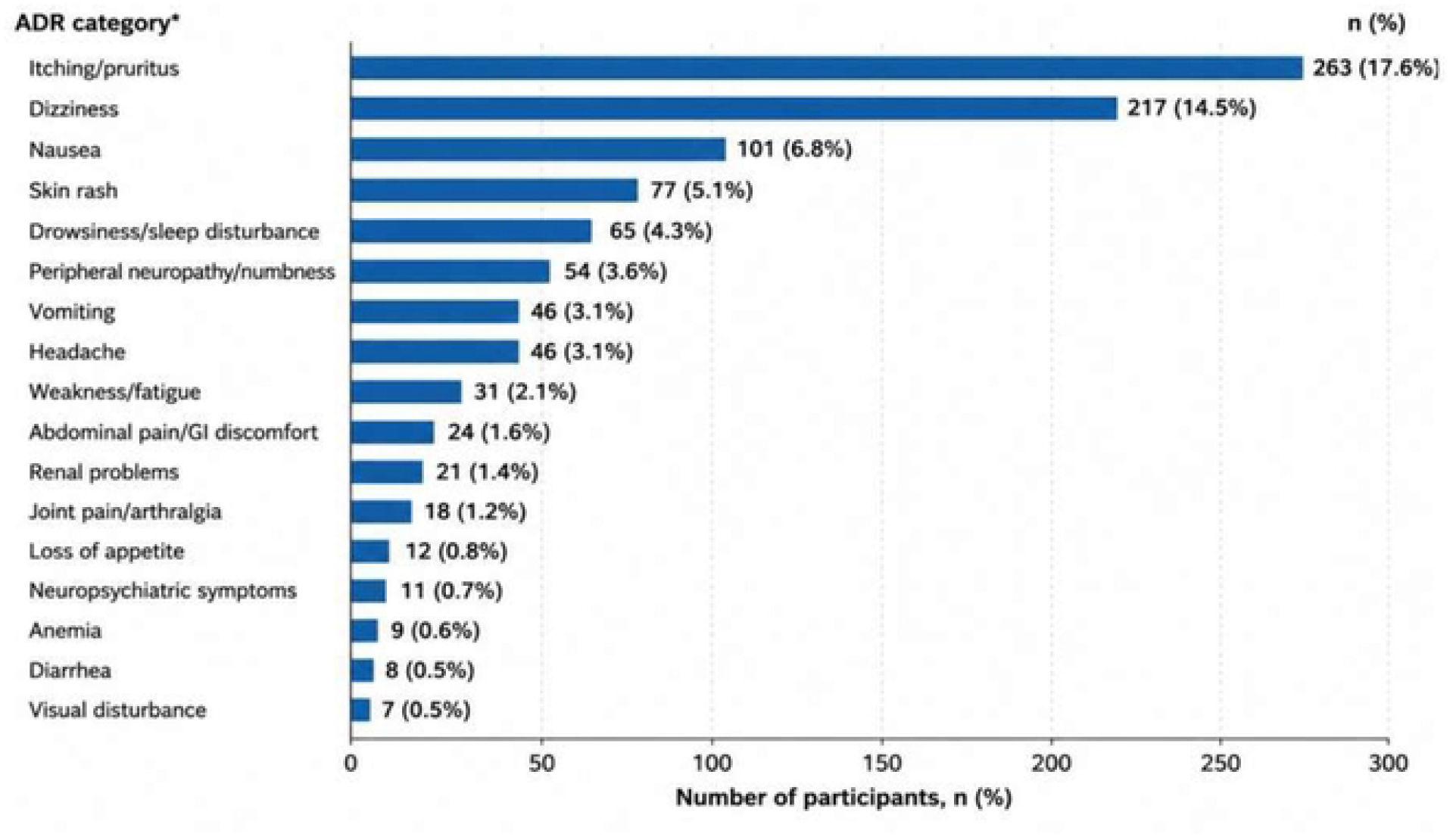
Distribution of reported ADR categories

### Association between advanced HIV disease and ADR occurrence

ADR prevalence was 38.8% among participants with AHD compared with 36.6% among participants without AHD. In univariate analysis, AHD was not significantly associated with ADR occurrence. In multivariable modified Poisson regression analysis, composite AHD was not independently associated with ADR occurrence after adjustment (aRR 0.95, 95% CI 0.82–1.11; p=0.537). However, the presence of comorbidity was strongly associated with ADR occurrence (aRR 1.66, 95% CI 1.42–1.93; p<0.001), while concomitant medication use was also independently associated with increased ADR risk (aRR 1.41, 95% CI 1.04–1.91; p=0.028) (Table 3).

**Table 3.** Univariate and multivariable modified Poisson regression analysis of factors associated with treatment-related ADRs.

| Variable | Crude RR (95% CI) | p-value | Adjusted RR (95% CI) | p-value |
| --- | --- | --- | --- | --- |
| **AHD** |  |  |  |  |
| No (Reference) | 1.00 | — | 1.00 | — |
| Yes | 1.06 (0.93–1.20) | 0.381 | 0.95 (0.82–1.11) | 0.537 |
| **Age group (years)** |  |  |  |  |
| 35–49 (Reference) | 1.00 | — | 1.00 | — |
| <18 | 0.71 (0.46–1.10) | 0.126 | 0.62 (0.37–1.03) | 0.065 |
| 18–34 | 1.08 (0.90–1.29) | 0.404 | 1.13 (0.93–1.37) | 0.224 |
| ≥50 | 0.95 (0.82–1.10) | 0.494 | 0.98 (0.84–1.15) | 0.817 |
| **Sex** |  |  |  |  |
| Female (Reference) | 1.00 | — | 1.00 | — |
| Male | 0.98 (0.85–1.13) | 0.778 | 0.97 (0.83–1.14) | 0.747 |
| **BMI category** |  |  |  |  |
| Normal BMI (Reference) | 1.00 | — | 1.00 | — |
| Underweight | 1.29 (1.03–1.61) | 0.026 | 1.21 (0.93–1.57) | 0.159 |
| Overweight | 0.92 (0.78–1.08) | 0.309 | 0.90 (0.76–1.07) | 0.235 |
| Obese | 1.04 (0.87–1.24) | 0.681 | 1.02 (0.84–1.23) | 0.844 |
| **ART regimen** |  |  |  |  |
| Non-DTG (Reference) | 1.00 | — | 1.00 | — |
| DTG-based | 0.83 (0.66–1.03) | 0.091 | 0.79 (0.61–1.03) | 0.087 |
| **Clinical characteristics** |  |  |  |  |
| **Presence of comorbidity** |  |  |  |  |
| None (Reference) | 1.00 | — | 1.00 | — |
| Yes | 1.58 (1.37–1.82) | <0.001 | 1.66 (1.42–1.93) | <0.001 |
| **Concomitant medication/OI prophylaxis** |  |  |  |  |
| None (Reference) | 1.00 | — | 1.00 | — |
| Yes | 1.48 (1.11–1.98) | 0.008 | 1.41 (1.04–1.91) | 0.028 |

### Sensitivity analysis

In adjusted sensitivity analyses, baseline CD4 count <200 cells/mm^3^ was independently associated with ADR occurrence (aRR 1.20, 95% CI 1.02–1.41; p=0.032), while WHO stage III/IV disease was not associated with ADR occurrence (aRR 1.01, 95% CI 0.86–1.18; p=0.918). Comorbidity remained strongly associated with ADR occurrence (aRR 1.71, 95% CI 1.44–2.03; p<0.001) (Table 4).

**Table 4.** Univariate and multivariable modified Poisson regression sensitivity analysis of advanced HIV disease components associated with treatment-related ADRs.

| Variable | Crude RR (95% CI) | p-value | Adjusted RR (95% CI) | p-value |
| --- | --- | --- | --- | --- |
| **Immunologic and clinical HIV disease markers** |  |  |  |  |
| CD4 ≥200 cells/mm<sup>3</sup> (Reference) | 1.00 | — | 1.00 | — |
| CD4 <200 cells/mm<sup>3</sup> | 1.29 (1.11–1.50) | 0.001 | 1.20 (1.02–1.41) | 0.032 |
| **WHO stage** |  |  |  |  |
| Stage I/II (Reference) | 1.00 | — | 1.00 | — |
| Stage III/IV | 1.08 (0.93–1.25) | 0.304 | 1.01 (0.86–1.18) | 0.918 |
| **Presence of comorbidity** |  |  |  |  |
| None (Reference) | 1.00 | — | 1.00 | — |
| Yes | 1.62 (1.40–1.87) | <0.001 | 1.71 (1.44–2.03) | <0.001 |
| **Concomitant medication/OI prophylaxis** |  |  |  |  |
| None (Reference) | 1.00 | — | 1.00 | — |
| Yes | 1.39 (1.05–1.84) | 0.021 | 1.31 (0.94–1.84) | 0.112 |
| **Treatment-related variables** |  |  |  |  |
| Non-DTG regimen (Reference) | 1.00 | — | 1.00 | — |
| DTG-based regimen | 0.81 (0.64–1.04) | 0.098 | 0.74 (0.57–0.96) | 0.024 |

## Discussion

This multicenter cross-sectional study among people living with HIV receiving ART in Dar es Salaam, Tanzania demonstrated a persistently high burden of AHD, with nearly two-thirds of participants (63.7%) meeting operational WHO-based criteria for AHD. ART-related adverse drug reactions (ADRs) were also common, affecting 38.0% of participants. While composite AHD was not independently associated with ADR occurrence, severe immunosuppression defined by baseline CD4 count <200 cells/mm^3^, comorbidity burden, and concomitant medication exposure were independently associated with increased ADR risk. These findings suggest that immunologic severity and broader clinical complexity may be more informative markers associated with ART-related toxicity than composite syndromic AHD classification alone.

The observed prevalence of AHD indicates a substantial and persistent burden of severe HIV disease despite widespread ART availability. This estimate is considerably higher than the approximately 33.5% pooled prevalence reported in outpatient HIV care settings globally, but is broadly comparable to estimates reported from hospitalized populations and several sub-Saharan African cohorts [5–7]. The high prevalence observed in our study may reflect several contextual factors. First, the study population included a large proportion of treatment-experienced individuals, suggesting that advanced disease in this setting may reflect not only delayed HIV diagnosis and late presentation to care, but also treatment interruption, poor retention, virologic failure, and immunologic deterioration after ART initiation [8–10]. Second, inclusion of both advanced clinical staging and severe immunosuppression in the operational AHD definition may yield higher prevalence estimates than studies relying solely on CD4 thresholds [4,5]. Third, participating facilities were urban referral HIV clinics managing clinically complex patients, which may have enriched the cohort with individuals experiencing persistent or recurrent advanced disease. Collectively, these findings suggest that AHD in the contemporary ART era increasingly represents an ongoing programmatic and clinical care challenge rather than solely a presentation-stage phenomenon.

Older age and underweight nutritional status were associated with higher AHD prevalence. The association with older age is consistent with previous studies demonstrating delayed HIV diagnosis, lower testing uptake, and poorer healthcare engagement among older adults [8,9]. In addition, aging populations living with HIV increasingly experience multimorbidity, treatment fatigue, and cumulative treatment exposure, which may contribute to advanced disease vulnerability [9,10]. Underweight participants also showed modestly higher AHD prevalence, consistent with regional evidence linking undernutrition and HIV disease progression [7,14]. HIV-related chronic inflammation, opportunistic infections, reduced appetite, gastrointestinal dysfunction, and metabolic catabolism may contribute to weight loss, while undernutrition itself may impair immune recovery and increase susceptibility to advanced disease [14]. Because of this bidirectional relationship, low BMI may represent both a marker and consequence of severe HIV disease.

Participants receiving DTG-based ART regimens had lower prevalence of AHD compared with those receiving non-DTG regimens. Although DTG-based regimens are associated with improved virologic potency, higher resistance barriers, and better tolerability compared with older regimens [15,16], this finding should be interpreted cautiously. The cross-sectional study design precludes causal inference, and alternative explanations are likely. Participants remaining on non-DTG regimens may represent individuals with historical ART exposure, prior treatment failure, toxicity-related regimen limitations, or more advanced disease requiring alternative treatment approaches. Additionally, because DTG rollout occurred more recently within Tanzanian HIV treatment programs, ART regimen category may partly reflect treatment era effects and historical regimen exposure. Accordingly, the observed association likely reflects residual confounding, survivor bias, and confounding by indication rather than direct causal protection against AHD.

ART-related ADRs affected more than one-third of participants, confirming that treatment toxicity remains clinically relevant in routine HIV care settings. The most frequently reported ADRs were dermatologic and neuro-gastrointestinal symptoms, including itching/pruritus, dizziness, nausea, and skin rash. These findings are generally consistent with pharmacovigilance studies from sub-Saharan Africa reporting persistent mild-to-moderate ART-related adverse events despite transition toward better tolerated regimens [11,17]. However, the ADR prevalence observed in our study appears somewhat higher than estimates reported in some contemporary DTG-era cohorts [15,18]. Several factors may explain this difference. First, our study used a broader ADR definition incorporating both participant-reported and clinically documented events, which may have increased event capture compared with studies relying solely on formal pharmacovigilance reporting systems. Second, the substantial burden of comorbidity and concomitant medication exposure in our cohort may have increased symptom burden and attribution complexity. Third, inclusion of clinically heterogeneous treatment-experienced participants may have captured cumulative toxicity related to prior ART exposure and regimen transitions.

Contrary to our initial hypothesis, composite AHD was not independently associated with ADR occurrence after adjustment. Although this initially appears counterintuitive, the finding may reflect the heterogeneous nature of the composite AHD construct. WHO-based AHD combines distinct clinical and immunologic dimensions, including severe immunosuppression and advanced clinical syndromes, which may not share identical biologic pathways influencing medication toxicity risk [4,14]. WHO clinical staging reflects overall disease burden, but some stage-defining conditions may not directly alter drug metabolism, physiologic reserve, or susceptibility to medication-related toxicity. Consequently, composite AHD may function as a useful prognostic and public health classification while remaining relatively imprecise as a marker of ART-related toxicity risk.

This interpretation is supported by the sensitivity analyses, in which baseline CD4 count <200 cells/mm^3^ remained independently associated with ADR occurrence whereas WHO stage III/IV disease did not. Severe immunosuppression may increase vulnerability to toxicity through impaired physiologic reserve, chronic systemic inflammation, malnutrition, opportunistic infections, organ dysfunction, and greater exposure to adjunctive therapies [11,13,14]. In contrast, WHO clinical staging alone may encompass heterogeneous disease states with differing implications for medication-related toxicity. These findings therefore suggest that immunologic severity may be more informative for identifying patients at increased risk of ART-related ADRs than syndromic disease staging alone.

Comorbidity emerged as the strongest correlate of ADR occurrence. Participants with comorbid conditions had substantially higher ADR risk even after multivariable adjustment. This finding is clinically expected because comorbid disease may increase medication burden, alter organ function, complicate symptom attribution, and increase vulnerability to cumulative toxicity and drug–drug interactions [19,20]. Similar associations between multimorbidity and medication-related adverse outcomes have been reported in HIV and non-HIV chronic disease populations [19,20]. These findings reinforce the evolving reality that HIV care increasingly involves management of chronic multimorbidity and complex medication exposure rather than isolated HIV treatment alone.

Concomitant medication use was also associated with increased ADR occurrence, supporting the role of polypharmacy in HIV pharmacovigilance. Concurrent medication exposure may increase toxicity risk through pharmacokinetic interactions, overlapping adverse-effect profiles, and treatment complexity [13,20,21]. However, because concomitant medication use may also reflect underlying disease severity and clinical complexity, residual confounding remains possible. This association should therefore be interpreted cautiously and viewed primarily as a marker of complex clinical care requirements rather than direct evidence of medication causality.

These findings have several clinical and programmatic implications. First, the persistently high burden of AHD supports continued prioritization of WHO-recommended AHD screening and differentiated care packages, including CD4 testing, opportunistic infection screening, prophylaxis, and intensified clinical monitoring [4,5]. Second, the finding that severe immunosuppression rather than composite AHD was associated with ADR occurrence suggests that immunologic risk stratification may be more informative for pharmacovigilance than syndromic staging alone. Third, the strong associations between comorbidity, concomitant medication use, and ADR occurrence underscore the importance of integrating medication review, interaction assessment, and broader chronic disease management into routine HIV care, particularly in aging populations increasingly affected by multimorbidity and polypharmacy [19– 21].

This study has several strengths. It included a large multicenter cohort drawn from routine HIV care settings, enhancing relevance to real-world clinical practice. Application of an operational WHO-based AHD definition improved comparability with existing literature. Use of modified Poisson regression allowed direct estimation of risk ratios for common outcomes, improving interpretability. In addition, sensitivity analyses separating composite AHD into individual clinical and immunologic components strengthened interpretation of the observed associations.

Several limitations should be acknowledged. First, the cross-sectional design precludes causal inference and limits assessment of temporal relationships between AHD markers and ADR occurrence. Second, ADR ascertainment included both participant-reported and clinically documented events, introducing potential recall bias, underreporting, and symptom misclassification. Third, WHO clinical staging reflected routinely documented clinical assessments obtained during HIV care and may not always have corresponded precisely with the timing of ADR occurrence. Fourth, some variables, particularly CD4 count and BMI, had missing observations, resulting in complete-case analyses that may have introduced selection bias. Fifth, information regarding cumulative ART duration and prior regimen exposure was incomplete, limiting assessment of historical treatment-related toxicity exposure. Finally, because the study was conducted in urban referral HIV care settings, generalizability to rural settings and hospitalized populations may be limited.

In conclusion, AHD remains highly prevalent among people living with HIV receiving ART in Tanzania despite widespread treatment availability. Although composite AHD was not independently associated with ART-related ADR occurrence, severe immunosuppression, comorbidity burden, and concomitant medication exposure were important correlates of toxicity.

These findings suggest that pharmacovigilance strategies should extend beyond syndromic HIV disease staging and incorporate immunologic severity, multimorbidity, and polypharmacy risk assessment within routine HIV care.

## Data Availability

The minimal data set is provided as other materials

